# Artificial intelligence directed development of a digital twin to measure soft tissue shift during head and neck surgery

**DOI:** 10.1101/2023.05.30.23290767

**Authors:** David Männle, Jan Pohlmann, Sara Monji-Azad, Jürgen Hesser, Nicole Rotter, Annette Affolter, Anne Lammert, Benedikt Kramer, Sonja Ludwig, Lena Huber, Claudia Scherl

## Abstract

**Introduction:** Digital twins derived from 3D scanning data were developed to measure soft tissue deformation in head and neck surgery by an artificial intelligence approach. This framework was applied suggesting feasibility of soft tissue shift detection as a hitherto unsolved problem.

**Methods:** In a pig head cadaver model 104 soft tissue resection had been performed. The surface of the removed soft tissue (RTP) and the corresponding resection cavity (RC) was scanned (N=416) to train an artificial intelligence (AI) with two different 3D object detectors (HoloLens 2; ArtecEva). An artificial tissue shift (TS) was created by changing the tissue temperature from 7,91±4,1°C to 36,37±1,28°C.

**Results:** Digital twins of RTP and RC in cold and warm conditions had been generated and volumes were calculated based on 3D surface meshes. Significant differences in number of vertices created by the different 3D scanners (HoloLens2 51313 vs. ArtecEva 21694, p<0.0001) hence result in differences in volume measurement of the RTC (p=0.0015). A significant TS could be induced by changing the temperature of the tissue of RC (p=0.0027) and RTP (p=<0.0001). RC showed more correlation in TS by heating than RTP with a volume increase of 3.1 μl or 9.09% (p=0.449).

**Conclusions:** Cadaver models are suitable for training a machine learning model for deformable registration through creation of a digital twin. Despite different point cloud densities, HoloLens and ArtecEva provide only slightly different estimates of volume. This means that both devices can be used for the task.TS can be simulated and measured by temperature change, in which RC and RTP react differently. This corresponds to the clinical behaviour of tumour and resection cavity during surgeries, which could be used for frozen section management and a range of other clinical applications.

## Introduction

Tissue shift (TS) is referred to as tissue displacements during soft tissue surgery. The tissue deforms after the wound opening due to its soft consistency and loss of tension. Consequently, anatomical landmarks can displace making orientation difficult. This effect has often been described in neurosurgery as brain shift and is considered a major source of error in neuro-navigation systems (1-3). Similar to neurosurgery, precise orientation is of immense importance in head and neck surgery, since many critical structures are located in very small space.

Therefore, navigation systems had been established. So far, these can only be used for the application of rigid bony structures, such as in sinus surgery (4). In visceral surgery marker-based tracking systems have been evaluated to aid in tumour resection and to compensate TS in soft tissue resections (5, 6). There are no marker-less navigation systems for head and neck surgery that compensate for soft tissue tracking. Resection of soft tissue tumours is a major field of head and neck surgery with tissue deformity causing difficulties. The study provides preliminary work for the development of a marker-free soft-tissue navigation system by determining TS using artificial intelligence (AI). Determining TS would have an enormous advantage for tumour resection itself and for handling frozen section procedure. Soft-tissue tumours could be resected better if orientation was improved by a navigation system. Furthermore, due to tissue deformation the exact spot for a re-resection after frozen section analysis is difficult to localize. Here, precise AI guided navigation can improve the safety, as areas to be re-resected can be detected in a better way.

Moreover, it is helpful to determine tissue deformation using AI in measuring and planning defect reconstruction with flaps. Due to TS, difficulties in fitting the flaps arise. The AI system developed in this study to determine TS could also help to gauge flap sizes more precisely in the future.

To the best authors’ knowledge, there is no study on AI measuring TS on volume changes of tumours and corresponding resection cavity. In order to obtain large amounts of data for AI training and the limited number of surgeries, the current study simulates tumour resections on pig cadaver heads to investigate a TS based on experimental volume changes. The study primarily intends to generate an AI system to register and measure TS. For that a simulation model for generating tissue shift was created.

## Material and Methods

### Animal cadavers and 3D scans

To conduct this experimental study 52 pig head cadavers (Schradi Frischfleisch GmbH, Mannheim) had been dissected. The animal cadaver model was approved by the Mannheim Veterinary Office (DE 08 222 1019 21). The pig heads were halved in the sagittal plane and stored at 7,91±4,1°C for immediate use or frozen at approximately -18°C for later processing. The pig head cadavers (PHC) were covered with surgical drape and tissue blocks at the parotid region down to the masseter muscle had been removed as a simulated tumour resection. Surgical drapes were marked next to the resection cavities and to the removed piece of tissue to make this region recognizable to the 3D cameras as “region of interest” (ROI) (**Fig 1**).

**Fig 1:**
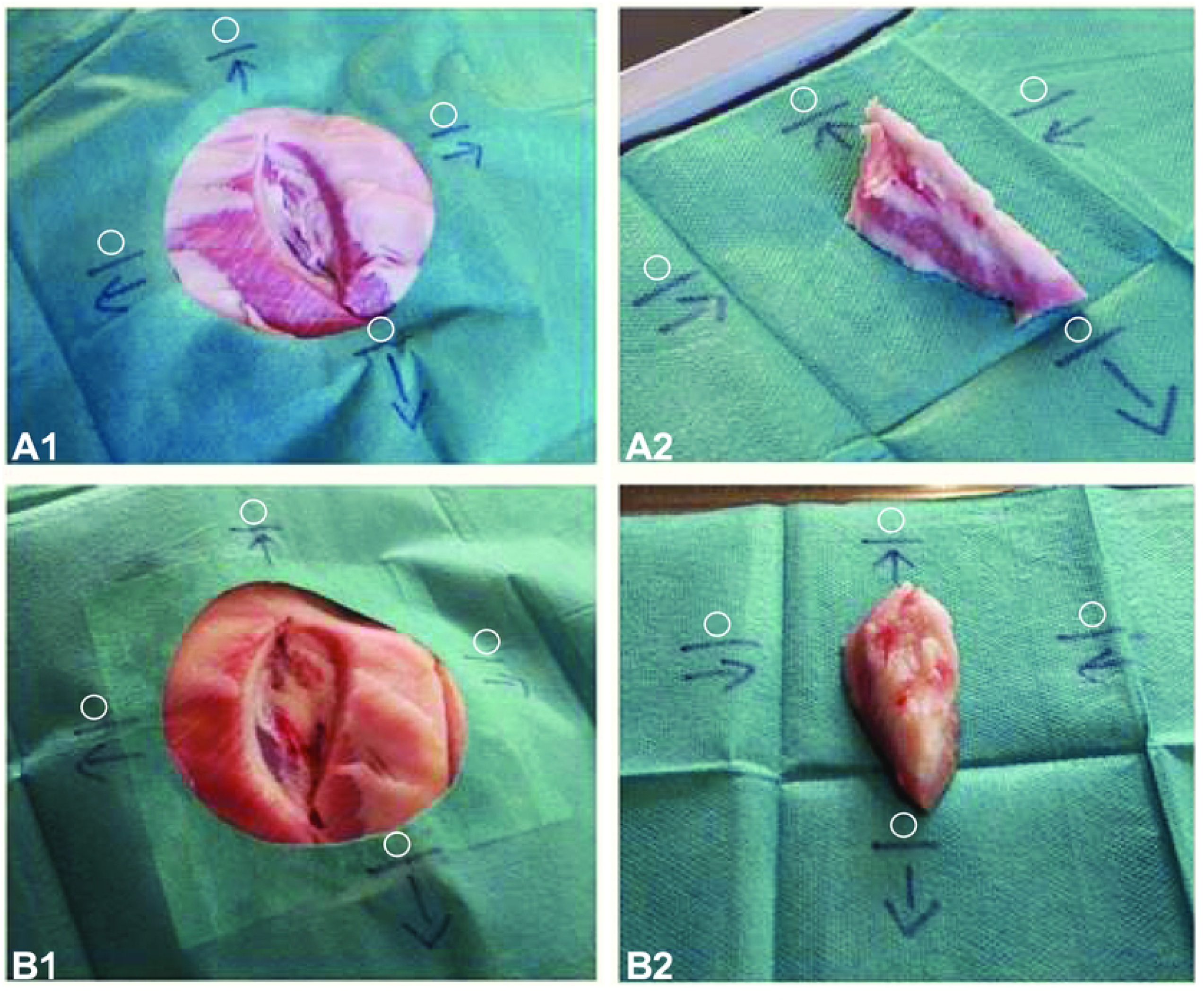
HMD images of pig head. Arrows and lines point to “cranial” (1), “caudal”(2), “rostral”(3), and “occipital”(4) direction to indicate the region of interest to the 3D-cameras and photogrammetry. A1: cold resection cavity; A2: cold removed tissue with basal side upwards; B1: warm resection cavity; A2: warm removed tissue with basal side upwards.

After resection the cavity and the resected tissue piece (RTP) are scanned with the 3D cameras (cold scan). Furthermore, images for photogrammetry are taken with the Head Mounted Display (see below). Tissue shift is simulated by heating. The shape of the tissue changes as a result of the temperature alteration, leading to a soft tissue displacement. In order to provoke a controlled and measurable tissue shift the RTP is placed back into the cavity, the PHC is covered with plastic sheets to minimize dehydration and warmed up to 36.37 ± 1.28 °C in a heat camber (*Binder GmbH, FD-53)* for 10 to 12 hours. The core temperature was measured continuously. Then immediately after warming the warm scans were performed.

To generate the raw data, scans were taken with a Head Mounted Display (HL2) (HoloLens 2©, Microsoft Corporation, Redmond, Washington, USA) and a 3D-object scanner ArtecEva (Artec3D, Luxembourg) (**Fig 2**). HoloLens and ArteEva have already been proven to be sufficient in the clinical-experimental use of head and neck surgery (7-9). Immediately before scanning core temperatures were measured using a piercing probe. With the HL2 approximately 15-20 images were taken from each angle in order to capture as much anatomical detail of the ROI as possible. To generate a detailed 3D mesh with the ArtecEva the camera is moved around the objects at a distance of 40 to 100 cm for 30-60 seconds.

**Fig 2:**
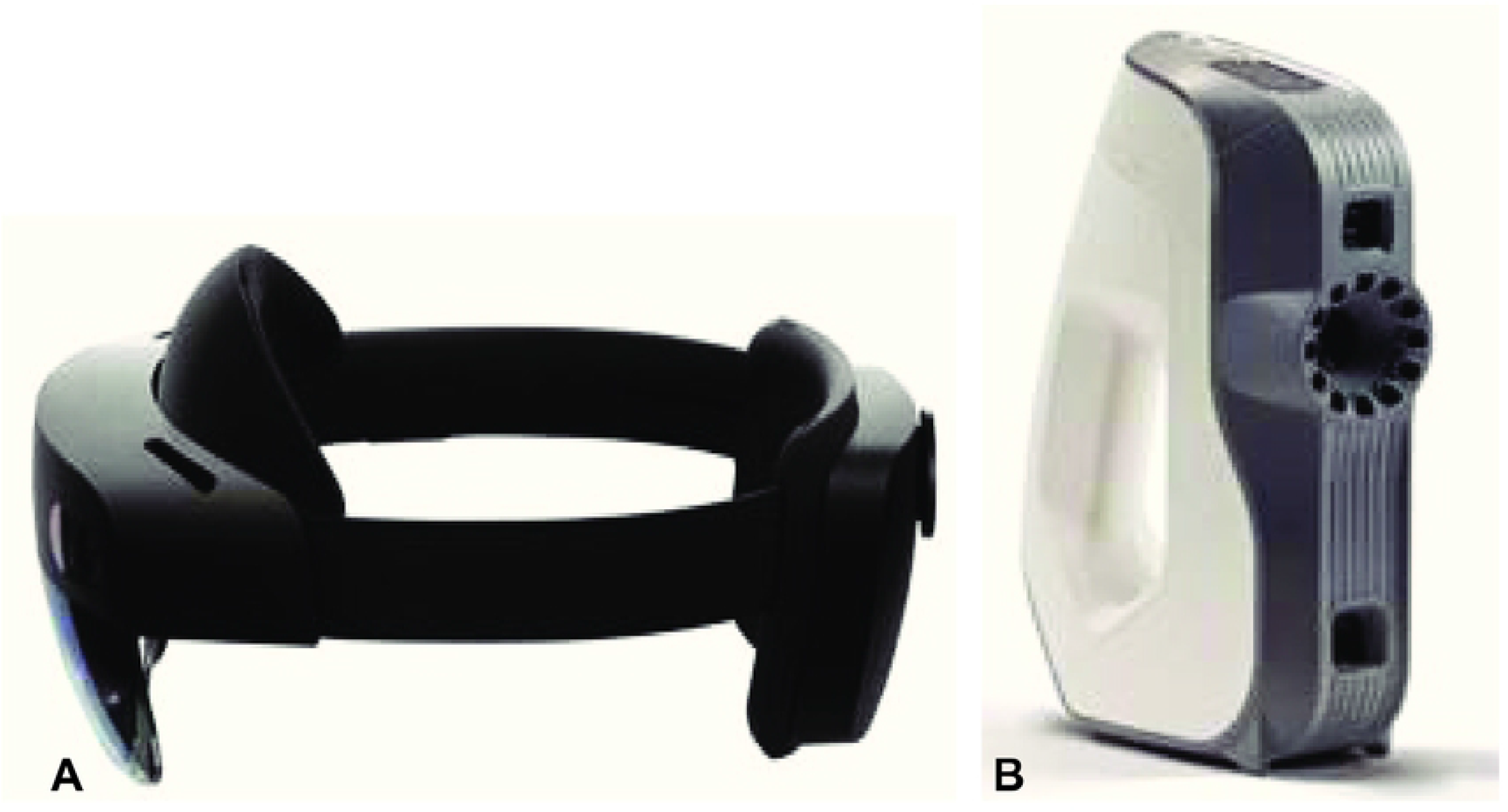
3D cameras. (a) Microsoft HoloLens 2© (Microsoft Corporation, Redmond, Washington, USA), (b) Artec Eva 3D object scanner (Artec3D, Luxembourg). The *HoloLens 2* is a mixed-reality headset that augments the vision by superimposing the reality with movable 3D holograms. 3D figures can be viewed from any perspective and virtually positioned anywhere in the user’s field of vision. Recordings are made with a camera integrated into the front part of the device in JPEG format (8 megapixels for still images and 1080 p30 pixels in video format). The 3D object scanner *ArtecEva* is hand-held 3D-camera being approved for medical use. It takes 16 pictures per second. The object to be scanned is illuminated alternately with a flashlight and a natural light. The 3D resolution is specified with an accuracy of 0.2mm.

### Data Generation

After the scans have been obtained, further processing takes place using photogrammetry. This process is done by using Meshroom software (Version 2021.1.0, Windows 10, Python 3.7.4) as a GUI for the Alice Vision Framework on a workstation with 16 Cores (AMD Ryzen 9 5950X) 64GB RAM and a single Nvidia 3080 Ti GPU. Mesh generation is done using Artec Studio 14 Professional, MeshLab 64bit, v2021.07, and Blender 2.93.4. Individual 2D images, which are created from different angles and are taken into account to form a 3D mesh. Then, Meshroom examines and compares the available input images, recognizes them, and is thus able to determine the different camera positions. Therefore, the 2D objects, depicted on the HL2 images in JPEG format, are converted into 3D figures.

### Post-Processing

To improve the quality of the 3D objects, some post-processing steps are done using MeshLab software. For instance, some of the 3D figures contain holes in their surface structure. Then the “Close Holes” function is used. Furthermore, during the creation and editing of the meshes, a total number is set to 300,000 vertices for the entire mesh. Since uneven distribution of the vertices on the 3D figures arise because some surfaces of the object are captured better than others during scanning, a MeshResampling node is added to Meshroom’s processing pipeline, and the “Simplification factor” setting is set to 1.0. It is worth mentioning that the development of the 3D meshes from the scans with the Artec Eva is carried out by the Artec Studio Professional 14 software, including the holes closing and ROI segmentation. During 3D mesh generation and post-processing steps, a 3D figure of each cavity and of the RTP is generated (**Fig 3**). The 3D figures created with Meshroom from the HL2 images are provided by the software in OBJ format. In contrast to the ArtecEva meshes, this format does not provide metric information such as the length and 3D object volume. However, for subsequent calculation of the volume of the resection cavity and the RTP the dimensionless sizes of the OBJ format have to be adapted to the real size. Additionally, an exact adjustment of the scales of the meshes from HL2 and Artec Eva images is necessary since the Meshroom software generates very small meshes compared to the real size of the objects. The metric information provided in the Artec meshes is considered the basis for calculating the scaling factor. Therefore, the length of the cranial line of the marks (**Fig 1**) is precisely measured three times in the MeshLab software and shown as “l_target_” in millimetres. Subsequently, the same measurement method is applied to the corresponding mesh from the HL2 recordings. The mean value is again calculated from the three measurements and shown as “l_source_”, which is dimensionless. Consequently, the scaling factor *f* is calculated using the following equation:

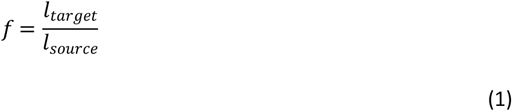

**Fig 3:**
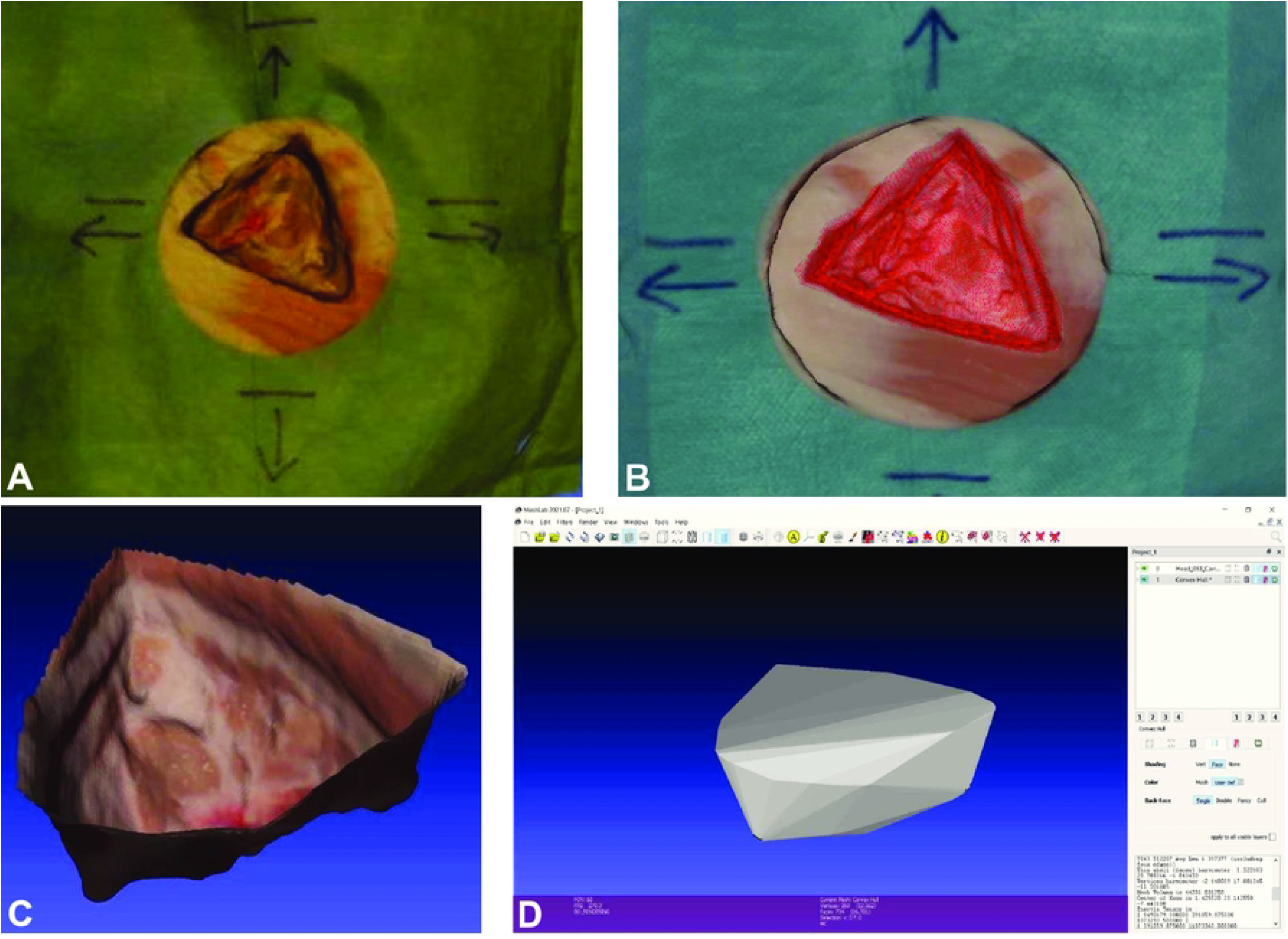
Segmentation of meshes with MeshLab of the cold resection cavity. A: Digital twin of HoloLens images; B: Digital twin of ArtecEva images; C: Completed segmentation of the ArtecEva mesh; D: View of the ArtecEva mesh after marking the volume of the ROI with the Convex Hull function and calculating it using Compute Geometric Measures

With the calculated scaling factors, we are able to adjust the dimensionless sizes of the HL2 meshes to the real scales.

### Segmentation and Volume

Segmenting the resection cavity and the RTP meshes is done in MeshLab. The “Draw Polyline” tool is used to digitally cut out the ROI from the rest of the mesh. To this end, the edges of the resection cavity and the RTP piece are marked out with the polyline by repeated mouse clicks. This is followed by an inversion of this marking to the portions outside the ROI, which will be deleted at the end. In this way, the relevant measurement area is separated from its external environment leaving only the resected cavity and the RTP for subsequent evaluation. The process of cavity and tumour segmentation to create a digital twin is shown in **Fig 3**. The volumes of the resection cavity and the RTP is measured in mm^3^ and also calculated using the MeshLab software. The output is shown in **Fig 3D**.

### Statistics

The Wilcoxon-signed-ranks test is used to compare the captured data using ArtecEva and HL2, with respect to the number of vertices in the ROI and the volume values of the cavity and of the RTP. Differences in volume as a function of temperature was also matched employing the Wilcoxon-signed-ranks test. This examines whether the central tendencies of two dependent paired samples differ. All analyses are reported with p-value, median, standard deviation (SD), minimum, maximum, range (minimum-maximum), and 95% confidence interval (CI). A result with a p-value < 0.05 is considered statistically significant. GraphPad PRISM, Version 9, 2020 was used for statistical analysis. Data were included in the final data aggregation (S1 appendix).

## Results

The collected data consist of temperature (°C), vertices, and volume amount (μl) which are provided both for cavity and RTP in cold and warm conditions. A total of 416 data sets from 104 halved pigs were evaluated. Due to insufficient quality problems of some meshes and some 3D objects 31 data sets were excluded from the present study as the ascertainment of vertices for determination the volumes were impossible, leaving 385 eligible cases for analysis. In this regard, 208 scans of cold tissue and 208 scans of warm tissue were unattainable.

### Vertices and volume depending on the capture device

To analyse the number of vertices in the ROI, 193 pairs of values of the meshes from HL2 and ArtecEva are used. The data which is examined include the segmented resection cavity and RTP of both, cold and warm, captures. As shown in **Fig 4A**, the HL2 generates a significantly higher number of vertices after processing the meshes compared to the ArtecEva camera. With a value of 51313 vertices, the HL2 meshes have more than twice the span in comparison to the 21694 vertices of the Artec 3D objects. The median and standard deviation of the vertices of the HL2 and the Artec Eva meshes are 12928 ± 6504 (range: 1145 to 52458, 95%-CI: 12344-14005) and 10158 ± 3079 (range: 1486 to 23180, 95%-CI: 9424-10632), respectively (p<0.0001).

**Fig 4:**
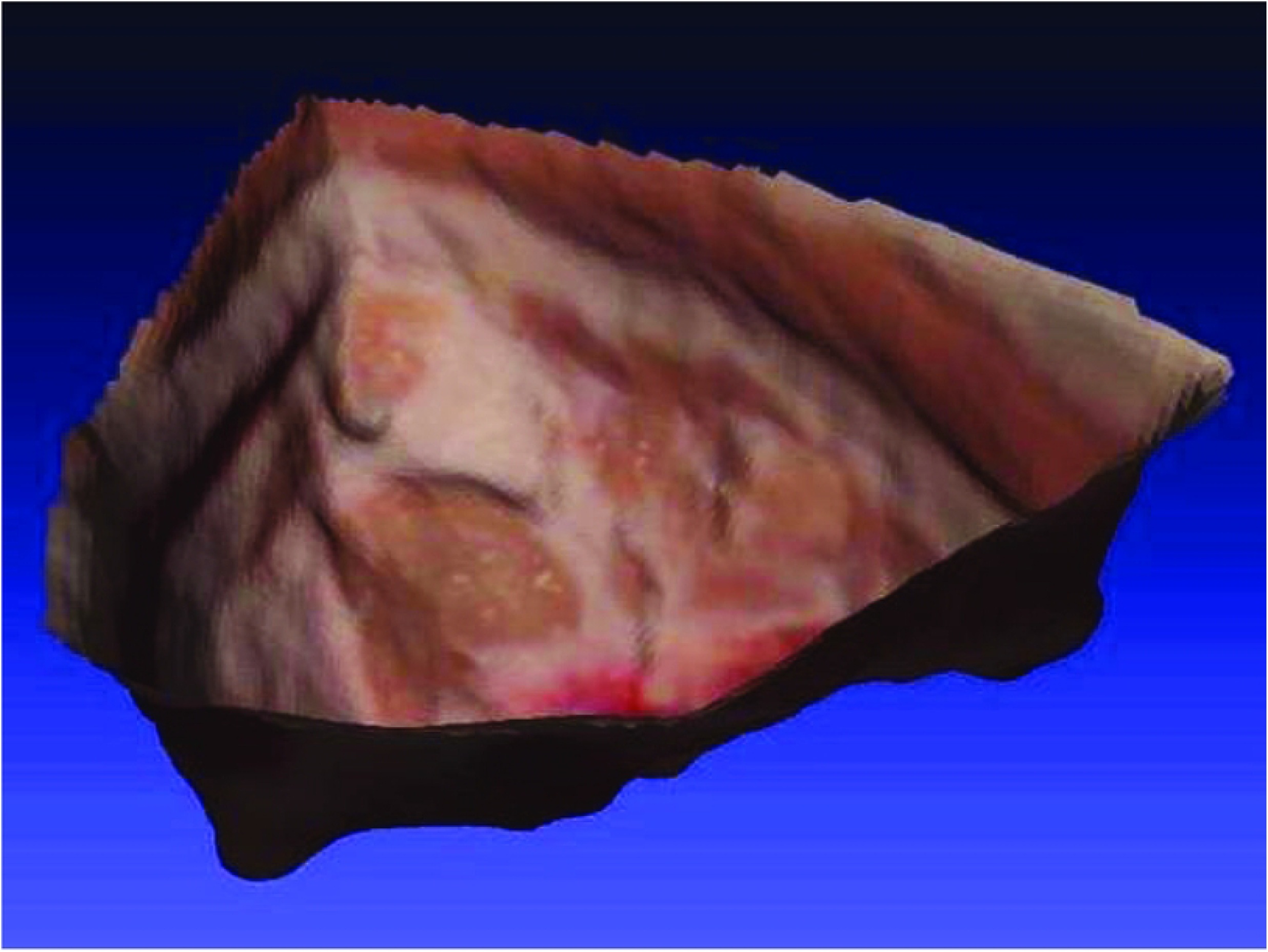
Vertices and volumes depending on the recording device. A: *Vertices* of resection cavity and RTP recorded either with HL2 or ArtecEva; B: Comparison of *volumes (μl)* depending on recording device for either cavity or RTP. HL2 = HoloLens, Artec = ArtecEva 3D object camera, RTP = resected tissue piece. Significance determined with Wilcoxon-signed-ranks test, p<0.0001 = highly significant***, p<0.01 very significant**, p<0.05 significant, ns = not significant.

Details of the different cameras regarding the volume (μl) of resection cavity and RTP are depicted in **Fig 4B**. Deeming the resection cavity, there was no significant difference between the HL2 (median ± SD: 35849 μl ± 17482 μl, range: 15814 μl to 119618 μl, 95%-CI: 30648-40188) and the ArtecEva (median ± SD: 36104 μl ± 17656 μl, range: 1253 μl -118037 μl, 95%-CI: 31370-38815) p=0.9116. Analysing the RTP volume the following significant differences are found: HL2 (median ± SD: 29425 μl ± 13636 μl, range: 13226 μl to 86991 μl, 95%-CI: 26217-33887) and ArtecEva (median ± SD: 31723 μl ± 14114 μl, range: 13696 μl to 92449 μl, 95%-CI: 26504-35296), (p=0.0015).

### Volume as a function of tissue and temperature

The following volume determinations refer to the data collected with the ArtecEva, as this device is already approved for medical applications. Significant differences in volume can be seen in **Fig 5A** comparing the cavity and the RTP for warm and cold temperatures. In the cold state (7-8°C) volume of the resection cavity and the RTP reveal significant differences (p=0.0449). The volume of the cavity revealed a median volume of 34074 μl ± 16970 μl (range 15373 μl to 118037 μl, 95%-CI: 28913-37990) and the RTP of 33318 μl ± 14524 μl (range 16485 μl to 92449 μl, 95% CI: 27105-37548). Comparing cavity and RTP in the warm state (36.37 ± 1.28°C), median volume of the cavity is 37173 μl ± 18496 μl, (range: 12531 μl to 117872 μl, 95%-CI: 29906-45620). In contrast, the warm RTP exhibited a median volume of 31199 μl ± 13501 μl (range 13696 μl to 85907 μl, 95%-CI: 23943-35576), p ≤ 0.0001.

**Fig 5:**
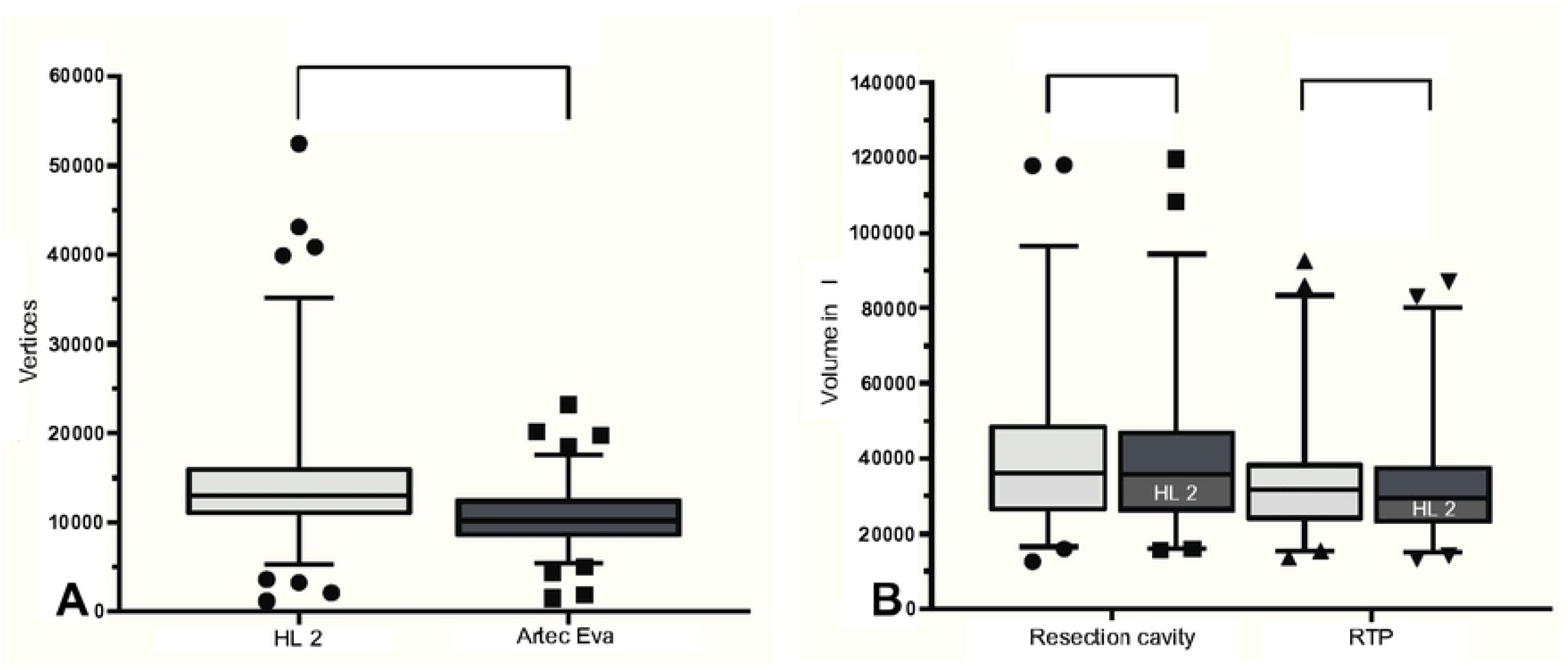
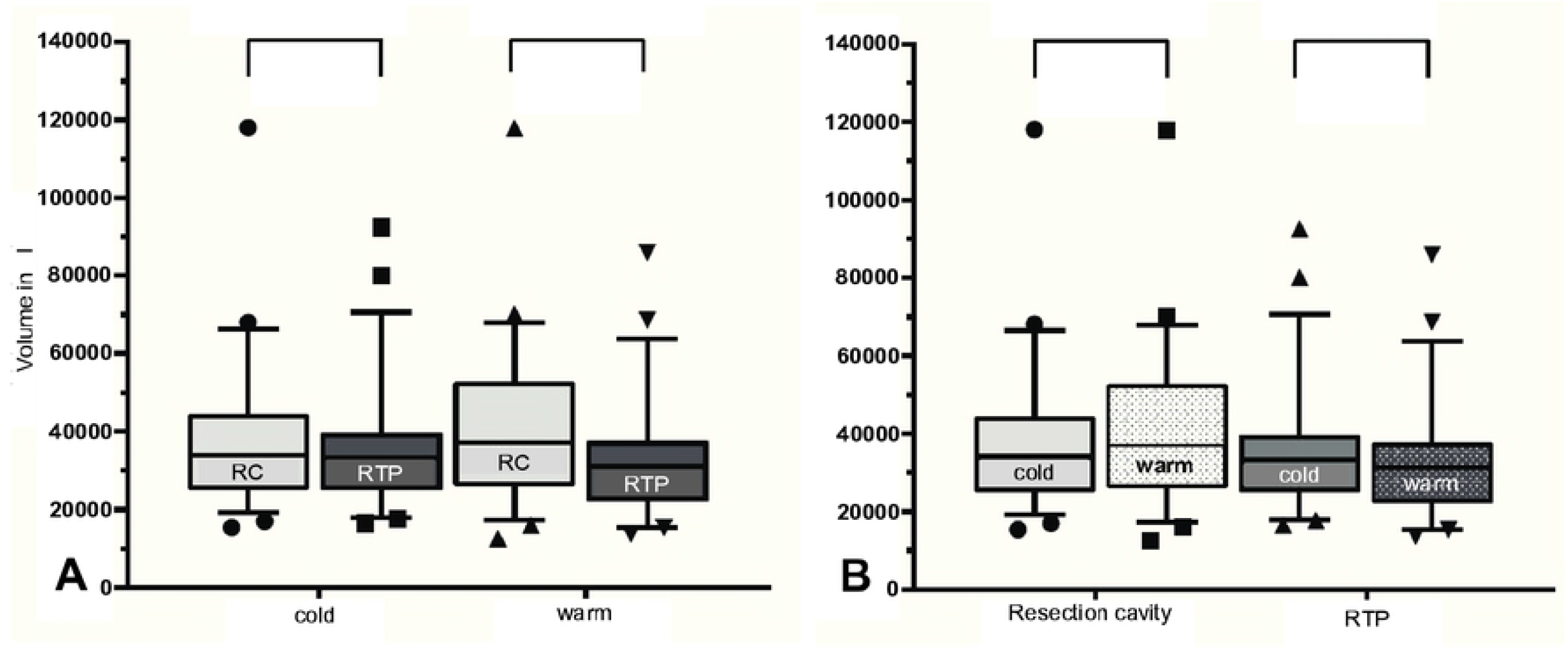
Volume as a function of tissue type and temperature. A: Volume as a function of tissue type: resection cavity (RC) and resected tissue piece (RTP) for cold and warm scans; B: Volume comparing cold (7-8°C) and warm (36.37 ± 1.28 °C) temperature of RC and RTP. RC = resection cavity, RTP = resected tissue piece. Significance determined with Wilcoxon-signed-ranks test, p<0.0001 = highly significant***, p<0.01 very significant**, p<0.05 significant, ns = not significant.

The boxplots of the volume of the resection cavity and RTP before and after heating of the tissue are shown in **Fig 5B**. The resection cavities increase the volume significantly by a median of 3.1 μl from 34.1 μl in the cold state to 37.2 μl after heating (p=0.0027). A highly significant decrease in RTP volume by median of 2.1 μl from 33.3 μl in the cold state to 31.2 μl after heating was measured (p<0.0001).

## Discussion

So far, no satisfactory method for measuring TS has been found. Numerous authors, mostly neurosurgeons, report on intraoperative imaging to determine TS during surgery (10-12). All methods are very time-consuming, costly and error-prone. With our AI based pilot investigation we describe a completely new approach. We were able to develop a pig carcass model on which soft tissue resections were performed. Tissue deformations were induced by controlled temperature changes. Hundreds of tissue scans trained an AI, which then registered and measured tissue deformation.

Two modern scanning devices were used and compared, Microsoft HoloLens 2 and Artec Eva 3D object scanner. Although the ArtecEva generates fewer vertices, the volume of the resection cavity is captured equally by both devices (p=0.9116). A higher number of vertices is not equivalent to an increase in accuracy. Many vertices of the HL2 overlapped. While requiring computing power the superimposed vertices did not contribute to volume calculation.

A TS can be simulated by changing the temperature of the tissue. This is caused by gaping of the borders of the resection cavity, loss of tissue tension and partial by dehydration. There is a change in Volume after heating. Cold tissue is rigid, thus preventing tissue to deform. The RTP and the defect remain precisely fitting when cold. The gaping of the borders of the cavity simulates the intraoperative behaviour of resection cavities quite well. During surgery the tension of the surrounding tissue leads to an enlargement of the defect. Our investigations show that temperature changes are suitable to simulate this intraoperative tissue shift of the resection cavity. The RTP volume decreases after heating. This is due to changes in the elastic properties of the tissue, dehydration, and leakage of fat. Here, too, the clinically known phenomenon of tumour shrinkage after resection can be simulated well.

The simulation model presented here could be used for the following clinical problems: (1) *Frozen section management:* This is where the largest and most important application potential can be seen. When assigning intraoperative frozen section findings, shifts in landmarks can occur due to TS in the resection cavity. That’s why target deviation for subsequent re-resection could be up to 1 cm (13). As a result, in 78% of all re-resection samples there is no residual tumour found. This could mean that the corresponding site in the tumour bed for the positive or close margins was incorrectly localized and tumour cells might remain in the patient (14). Our method can help to measure the TS in order to better determine areas for subsequent re-resection to remove the tumour completely. This increases the oncological safety of frozen section-controlled tumour resections. (2) *Flap planning:* In head and neck surgery large defects have to be closed by tissue transfer (flaps). During surgery it is common to estimate the size of a flap before tumour removal by the size of the tumour. When planning to close the defect with a flap in that way, fitting difficulties of the flap are possible later on due to a TS on the resection cavity. The method shown in this report can be used to determine the size and volume of the defect intraoperatively in order to better determine the dimensions and volume of the flap. In addition, a subsequent flap shrinkage must be taken into account. It is recommended to include this shrinkage in the flap size planning beforehand (15, 16). Due to the possibility of precisely determining the volume of the resection cavity with the presented method, it could also help to calculate the amount of overcorrection. (3) *Soft tissue navigation:* In soft tissue navigation inaccuracies occur due to TS. Marker-based tracking systems should help to compensate TS, which is also not error-free (5). As a basis for the development of soft tissue navigation without markers, we present a completely new AI-based approach. Similar to the method by Pfeiffer M et al for liver surgery (17) we developed a system to record tissue deformation in real-time, which could be an enormous orientation aid, especially in head and neck surgery. Because, due to the anatomically very narrow conditions with many structures in a small space, it is difficult to find tumours and remove them in such a way that important structures are spared.

Based on the investigations, it shows that generating controlled TS is possible in an animal cadaver model and to measure it by volume differences. In future works, this methodology can be applied to real tumour resection during soft tissue surgery. For this, additional work is needed in training AI to augment model prediction and accuracy, which than will also recognize different tumour formations.

## Conclusions

In this study, we verified that cadaver models are suitable for training an AI through creation of big data. In the scanning process, the number of vertices depends on the recording device. The more vertices were generated, the more superimpositions of these occurred, which are not included in the volume calculation. TS can be simulated by temperature change mirroring a number of clinical phenomena, for which the presented method can be helpful in the future.

## Data Availability

All relevant data are within the manuscript and its Supporting Information files

## Acknowledgements

We would like to thank Johann Kern and Petra Prohaska for the provision of animal laboratory space including all necessary equipment and their assistance during resection and scanning process. Furthermore, we thank Dennis Feiler and Holger Ladewig (DFC SYSTEMS, Munich) for excellent technical support. The study was funded by the German Federal Ministry of Economic Affairs and Climate Action, Central Innovation Programme for small and medium-sized enterprises (SMEs); funding number: KK5044704CS0.

## Notes

Data Availability Statement All relevant data are within the paper and its Supporting Information files.

### Competing Interest Statement

The authors have declared no competing interest.

### Author Declarations

No ethics committee approval necessary

